# Trends in public awareness of sepsis in Australia from 2016 to 2025: a longitudinal analysis of four national surveys (Preliminary results)

**DOI:** 10.64898/2025.12.03.25341602

**Authors:** Ashwani Kumar, Brett Abbenbroek, Jacob Dye, Kelly Thompson, Luka Saxena, Simon Finfer, Naomi E. Hammond, Sepsis Australia

**Author notes:** Corresponding author The George Institute for Global Health, Level 8, Health Translation Hub, 55 Botany Street Randwick NSW 2031 Australia.

## Abstract

**Objectives and importance of study:** Sepsis is a time-critical life-threatening condition, with over 80,000 cases annually in Australia. With approximately 80% of sepsis cases arising in the community, public awareness is essential for early recognition, prompt treatment and reducing sepsis-related morbidity and mortality. This study aimed to assess and report changes in public awareness about sepsis over time in the context of awareness initiatives.

**Study type:** Longitudinal analysis of nationally representative surveys.

**Methods:** Sepsis Australia partnered with YouGov^®^ to develop, conduct, and administer four national surveys between 2016 and 2025 to assess public awareness of sepsis. Data collected from a representative sample of Australians aged ≥18 years included socio-demographic characteristics and measures of sepsis awareness and knowledge. Additional questions were progressively introduced on the link between sepsis and COVID-19, First Nations status, high-risk groups, post-sepsis syndrome (PSS), information sources, and perceived government support. Data were analysed descriptively, trends assessed using logistic regression, and subgroups (gender, age, residence, marital status, children [<18 years] at home, household income, and generation) compared using Chi-square tests with significance set at p<0.05. Comparisons were made across subgroups for each year, unless specified.

**Results:** The sample over the four surveys’, ranged between 1000 to 1232 respondents with similar demographic characteristics. Sepsis awareness increased significantly from 40% (2016) to 70% (2025; p<.001), but remained lower than for other conditions. Knowledge of symptoms; mortality (4%-6%), PSS (18%) and high-risk groups, such as pregnancy/postpartum (8%), was low. Women (46%-75% vs 34%-65% in men, p<.001), those aged ≥50 years (47%-83% vs 28%-47% aged 18-24 years, p=.01), those without a child (40%-75% vs 39%-62%, p<.001), non-capital city residents (43%-80% vs 38%-64%, p<.001 for 2022 and 2025), and unemployed respondents (46%-77% vs 35%-63%, p<.01) reported higher awareness. Differences in other subgroups were not significant. Social circle was the most common (34%) source of sepsis information and 46% respondents advocated for government support.

**Conclusion:** The Australian general public awareness of sepsis has increased in recent years, coinciding with initiatives undertaken by Sepsis Australia and other organisations, though direct causality cannot be inferred. Gaps persist in awareness among men, and younger adults, and knowledge of PSS and high-risk groups is limited. A comprehensive, targeted awareness campaign should be considered to improve public awareness of sepsis.

**Key points:**

- Sepsis mortality and morbidity is preventable through public awareness that promotes early recognition and triggers action to seek advice or emergency care.
- Sepsis awareness in Australia has increased significantly in recent years but remains lower than other conditions.
- Lower awareness of sepsis was reported in men, young adults, parents of children younger than 18 years, and in those who were employed.
- Knowledge of post-sepsis syndrome and risk factors for sepsis is low.
- A comprehensive public awareness is warranted to address the growing burden of sepsis in Australia.

## Introduction

Sepsis is a time critical life-threatening dysregulated host response to infection that leads to organ dysfunction, tissue damage and potentially death. It is a global health challenge and is the leading cause of preventable deaths globally. (1) Recent global and Australian estimates indicate a three-fold increase in the magnitude of sepsis burden. (2–4) As more than 80% of sepsis cases originate in the community, improving patient survival and outcomes depends on early recognition and timely medical care. (5, 6) Achieving this requires enhanced public knowledge and awareness of sepsis; however, public awareness of sepsis in many countries is poor. (7, 8)

In 2017, Australia co-sponsored a World Health Organisation (WHO) resolution on improving the prevention, diagnosis and clinical management of sepsis. The 70th World Health Assembly adopted the resolution (70.7), urging all 194 United Nations member states to take national action on sepsis. (1) Sepsis Australia, formerly the Australian Sepsis Network, was established in 2016 within the Critical Care Program of The George Institute for Global Health. (9) Despite a lack of dedicated program funding, Sepsis Australia has played a crucial role in coordinating national sepsis initiatives since its inception. Its first major initiative was a policy roundtable bringing together consumers, clinicians, researchers, government agencies, and professional societies to formulate the Stopping Sepsis National Action Plan (SSNAP). Launched in 2017, one of the SSNAP’s key strategic goals was to educate and increase sepsis awareness among both healthcare professionals and the public. Under the SSNAP strategic framework, Sepsis Australia has led and coordinated national initiatives across awareness, prevention, education, clinical quality improvement, research and consumer engagement. These include collaboration with the Australian Commission on Quality and Safety in Healthcare (ACSQHC) on social media awareness activities, annual World Sepsis Day campaigns, and development of the national Sepsis Clinical Care Standard (2022). (10, 11) A cornerstone of these efforts is the Sepsis Australia Consumer Partner and Advocacy Program (SACPAP), comprising a 48-member consumer leadership group representing sepsis survivors, their families and carers, and families bereaved by sepsis.

To evaluate the potential impact of these initiatives and guide future strategies, four national surveys were conducted between 2016 and 2025 to assess public awareness of sepsis, its attributes and consequences, and compare awareness with other major medical conditions such as heart attack, breast cancer and leukaemia. The first survey provided baseline data, followed by three surveys in 2020, 2022 and 2025 enabling longitudinal analysis of trends in public awareness of sepsis in Australia.

## Methods

On behalf of Sepsis Australia, four cross-sectional, national online surveys were conducted by YouGov Plc. (https://au.yougov.com), an international online research data and analytics group. Surveys were administered over one-week period in August 2016, July 2020, November 2022 and June 2025. In each round, a survey measuring awareness (12) was distributed to an unidentified and nationally representative sample of the Australians aged 18 years and older. Survey participation was voluntary and its completion implied participant consent for researchers to share and publish the summary data.

Respondents were distributed across all Australian states and territories, and participant samples were weighted by age, gender and region to be representative of the adult population of Australia using the latest population estimates from the Australian Bureau of Statistics (ABS) (https://www.abs.gov.au/).

### Questionnaire

All four surveys used an English-language questionnaire developed by Sepsis Australia. The 2016 survey questionnaire had eight questions, including three on demographic characteristics and five on sepsis awareness. In 2020, one additional question was included on the relationship between COVID-19 and sepsis. The 2022 survey was expanded to 11 questions, whereas the 2025 survey included five additional questions. Questions included demographics, and sepsis awareness questions with multiple choice or open-ended responses (Refer Supplementary File 1 for details).

### Data collection

Demographic data collected included age, gender, marital status, presence of children under 18 years in the household, employment status, household income and place of residence. First Nations status was captured in 2022 and 2025. Sepsis awareness data included level of awareness and other common medical conditions, knowledge of someone who had sepsis, perceived mortality, recall of symptoms and causes, awareness of the association between COVID-19 and sepsis (all except 2016 survey), and media coverage of sepsis in the previous 12 months (2022 and 2025 only). In the 2025 survey, additional data related to knowledge of post-sepsis syndrome (PSS), awareness of at-risk groups, and perception of adequacy of government support were collected. In addition, respondents with lived experience of sepsis were also asked about sources of information and details of lived experience of sepsis.

### Data management

Data were stored within secure servers hosted within designated secure data centres. Independent penetration tests are undertaken by YouGov^®^ on an annual basis, and systems and applications and continually scanned for vulnerabilities. Data encryption was used to secure data whilst in transit using AES256 encryption.

### Data analysis

De-identified survey data were analysed and reported using descriptive statistics as frequency with percentage. Data analysis was performed using Microsoft Excel (Microsoft Corporation 2018; https://office.microsoft.com/excel) and IBM SPSS Statistics 31.0.0.0. The subgroups used to report the survey results were age groups in years, gender, geographical location, household income, marital status, children younger than 18 years at home, First Nations status, and various generations (GenZ [1997-2009], Next Gen [18-34 years], Millennials [1981-1996], GenX [1965-1980], Baby Boomers [1946-1964], Silent [1918-1945]). Temporal trends in overall sepsis awareness were calculated using binary logistic regression with year (centred at 2016) as a continuous predictor with case weights equal to the aggregated counts per year derived from the survey numbers and reported percentages. Sepsis awareness was compared using Chi-square test in following subgroups: gender, age groups, state and territory, capital city residence, children younger than 18 years, marital status, household income, employment status, generation and First nation status. P values <.05 were considered as statistically significant. Other aspects of sepsis awareness are presented in descriptive manner. We used EQUATOR Checklist for Reporting of Survey Studies (CROSS) to report our results. (13)

## Results

### Demographic characteristics

The 2016, 2020, 2022, and 2025 surveys included 1000, 1006, 1131, and 1232 participants, respectively. Demographic characteristics of the participants were comparable across all four surveys (Table 1).

**Table 1.**
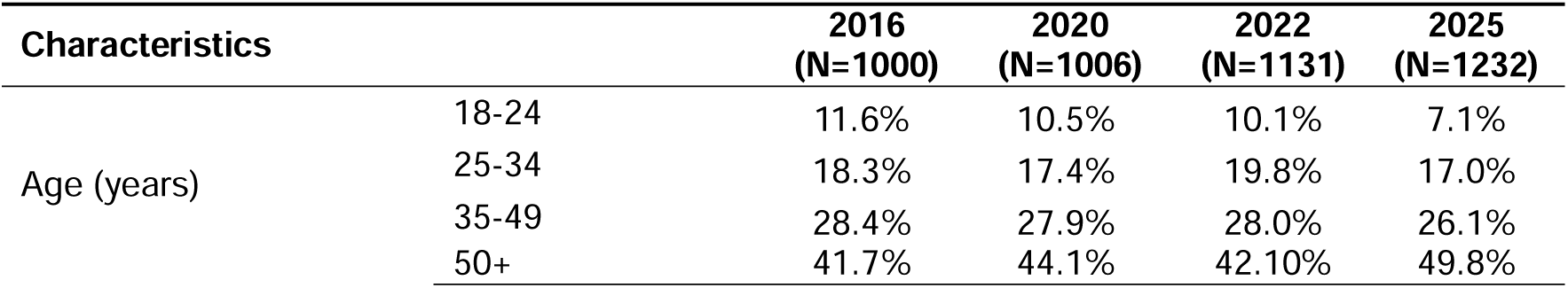
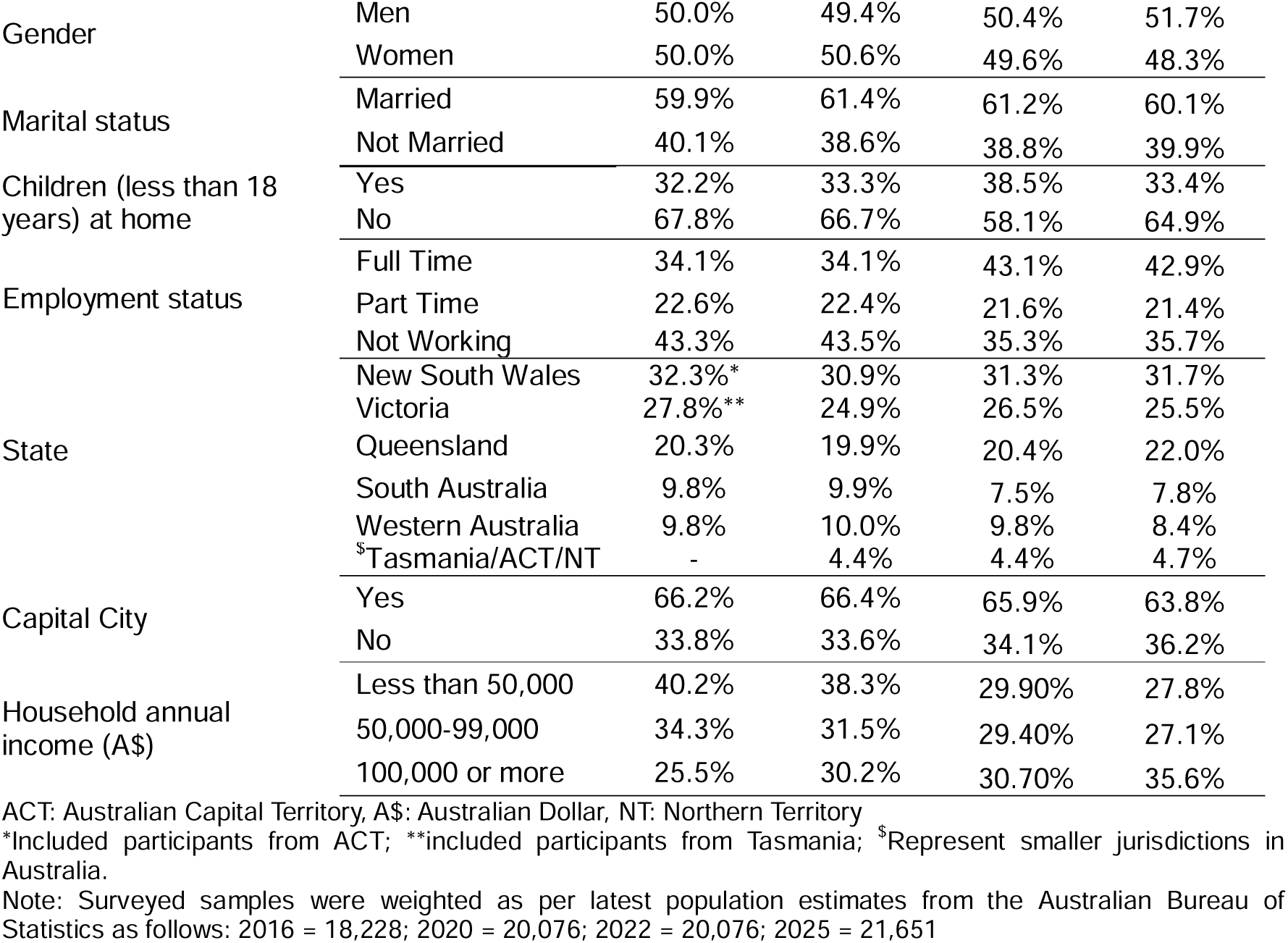
Demographic characteristics of the survey participants.

### Overall awareness of sepsis

A statistically significant increase in sepsis awareness was noted from 40% in 2016 to 70% in 2025 (p<.001). Compared to other medical conditions, such as breast cancer and heart attack, awareness of sepsis was still numerically lower. The largest increase occurred between the first and second surveys (19%) followed by between third and fourth surveys (9%) (Figure 1 and Table 2).

**Figure 1.**
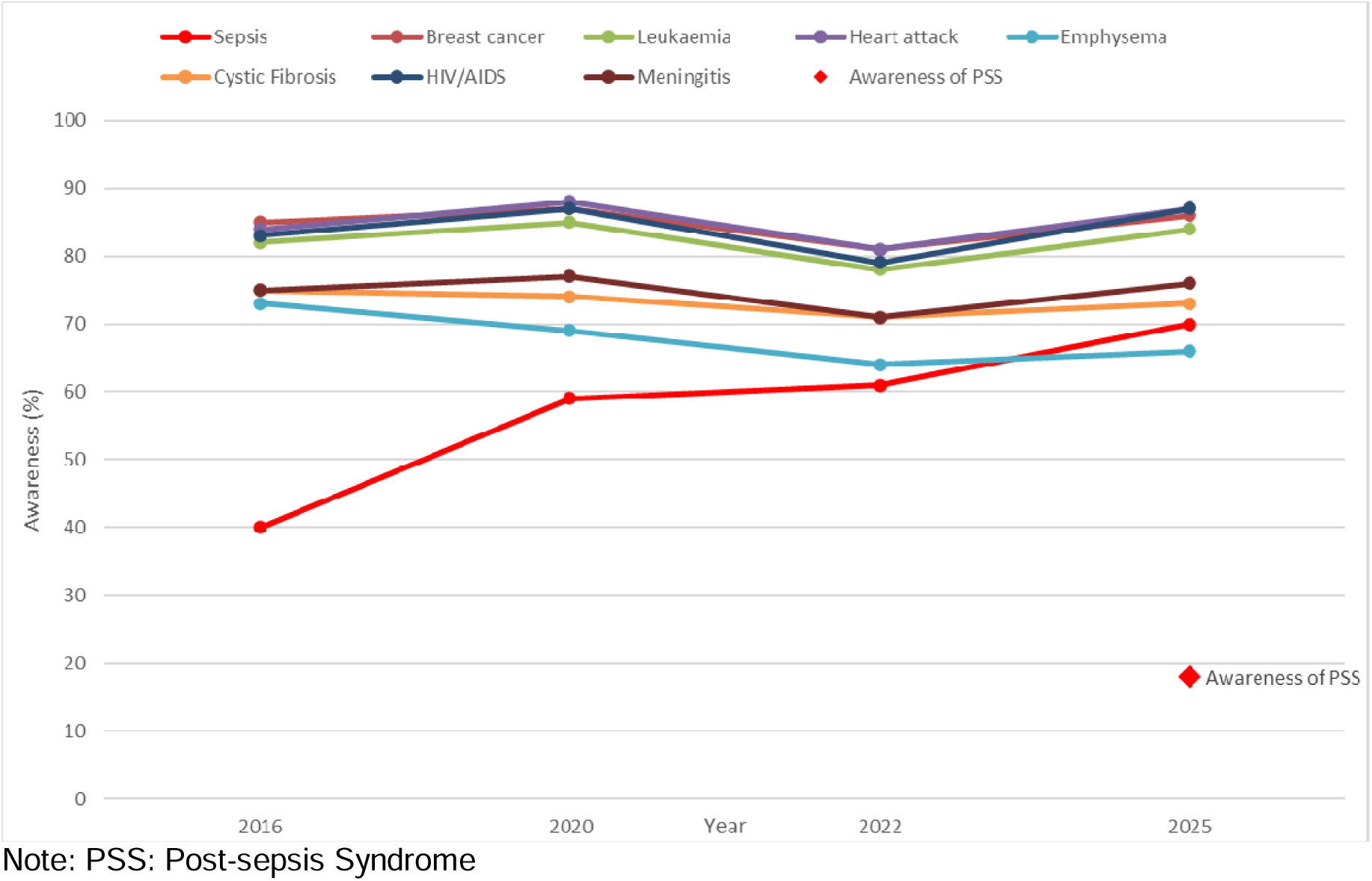
Awareness of sepsis and various medical conditions.

**Table 2.**
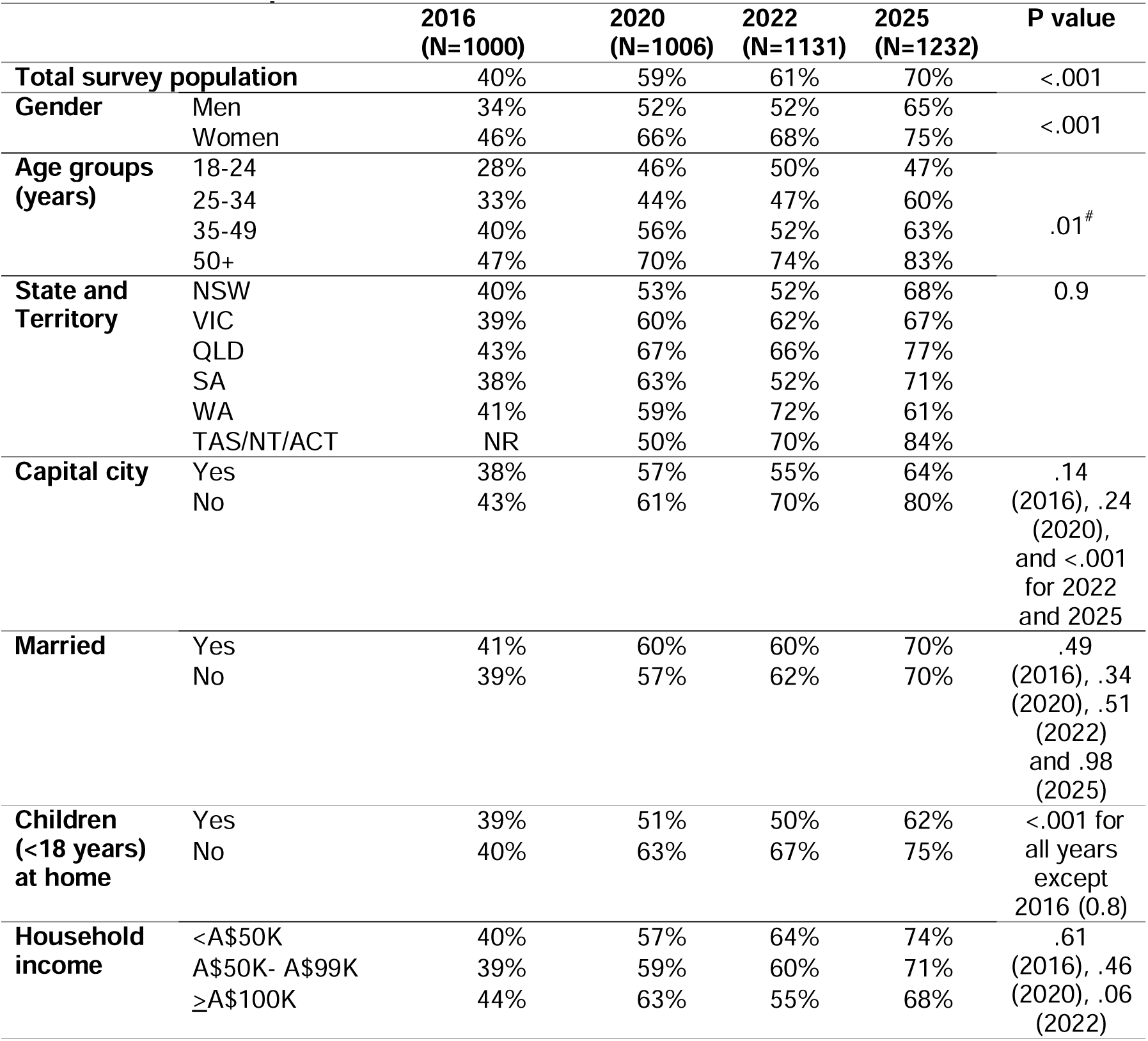
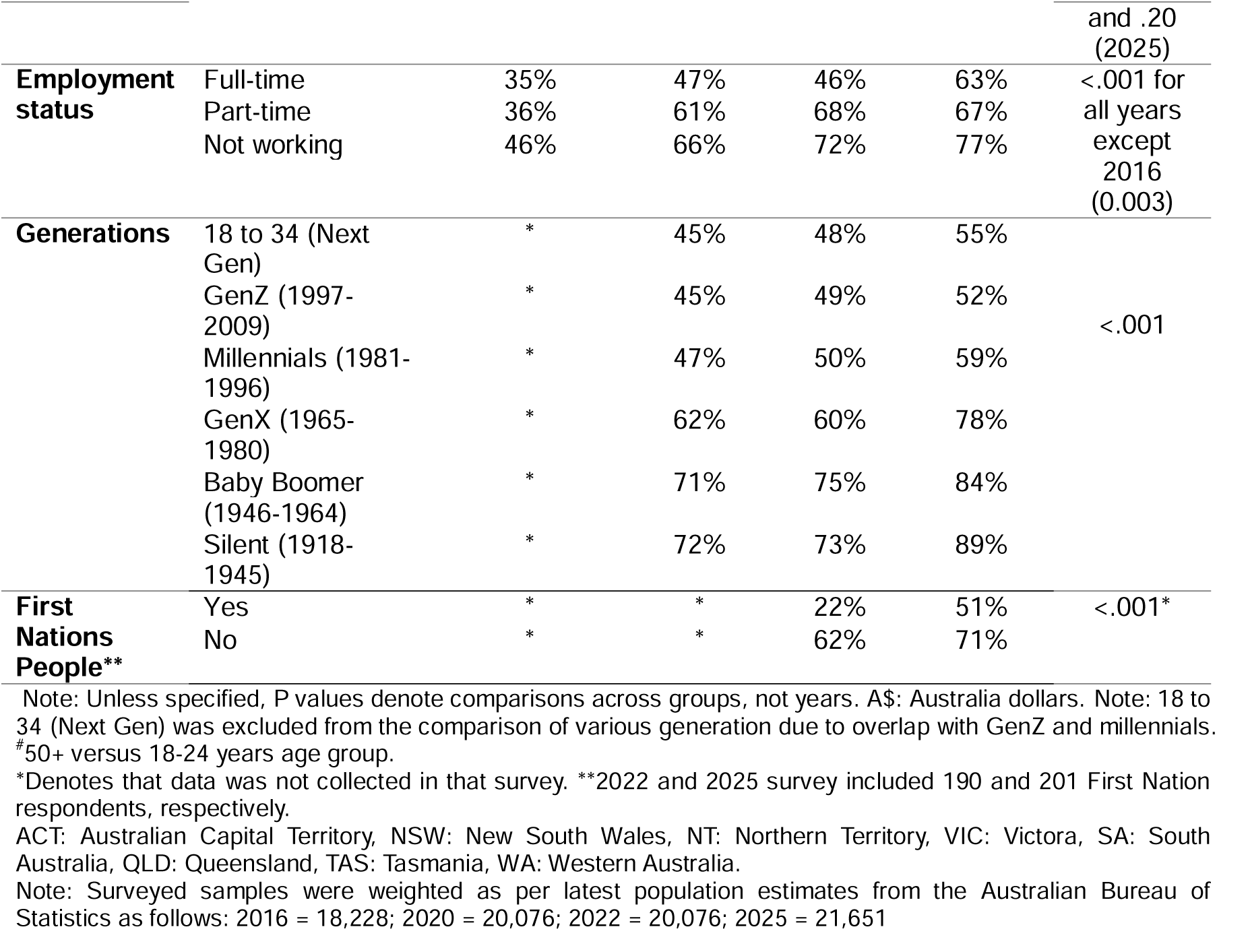
Overall sepsis awareness.

### Overall sepsis awareness in specific subgroups

Sepsis awareness increased among both men and women. However, women consistently reported significantly higher overall sepsis awareness (46%-75%) than men (34%-65%), all p<.001. Respondents aged 50 years and older demonstrated significantly higher awareness (47%-83%; p=.01) than those aged 18-24 years. Sepsis awareness differed significantly (p<.05) across states in all surveys, except 2016. Among capital city residents, awareness increased from 38% to 64% and among non-capital residents 43% to 80%, with differences being statistically significant in 2022 and 2025 (both p<.001).

Sepsis awareness was similar in married and unmarried respondents, whereas parents of children under 18 years reported significantly lower awareness (p<.001), except in 2016. No statistically significant differences were observed between income groups across surveys. Unemployed respondents consistently demonstrated significantly higher awareness (all p<.01) across surveys compared with employed respondents. All generations showed an increase in awareness, with the highest awareness observed in the silent generation (72%-89%), followed by Baby Boomers (71%-84%), and the lowest in GenZ (45%-52%). Differences in awareness between generations were statistically significant (p<.001) across all surveys (Table 2).

In 2022, sepsis awareness was significantly lower among First Nations respondents compared to other Australians (22% vs 62%, p<.001). By 2025, awareness among First Nations people increased to 51%, narrowing the gap with other Australians (71%), though the difference remained statistically significant (p<.001) (Table 2 and Figure S2).

### Sepsis awareness and key milestones

Figure 2 illustrates the timeline of key activities undertaken by Sepsis Australia and other organisations including the ACSQHC, to improve sepsis awareness during the period of four surveys, alongside the trend in sepsis awareness. In total 21 activities were undertaken between 2016 and 2025, with more than two-thirds occurring between 2021 and 2025.

**Figure 2.**
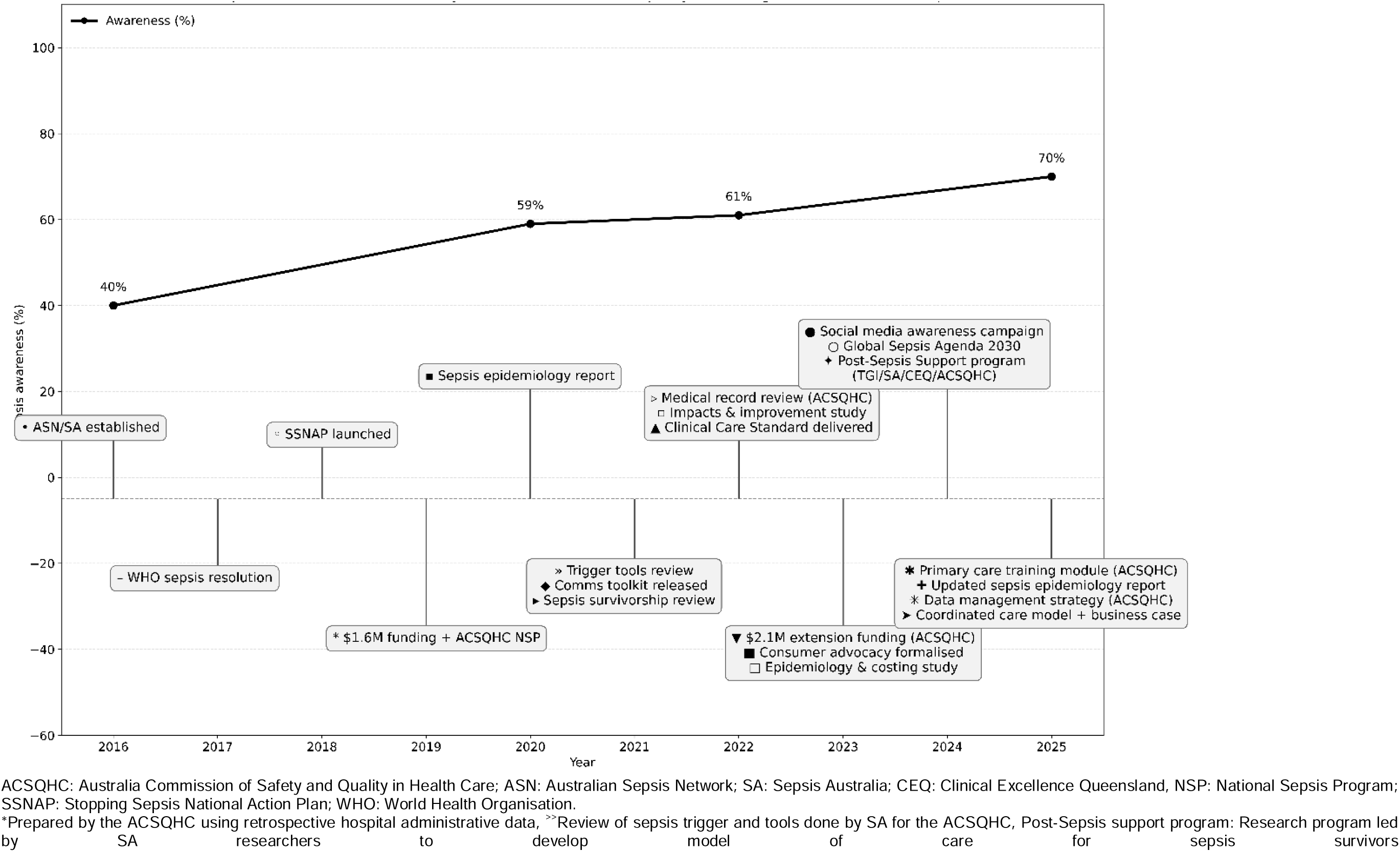
Sepsis awareness trend and key milestones.

### Other aspects of sepsis awareness

Recognition of fever, chills and sweating as a sepsis symptom cluster increased from 14% (2016) to 36% (2025). Other infection was the most frequently reported cause of sepsis (11%-32%; Table S1). Awareness of the sepsis-COVID-19 link peaked in 2022. In 2025, only 31% respondents identify any at-risk groups for sepsis, most commonly older age (41%) and immunodeficient states (40%), while pregnancy/ postpartum remained least recognised (8%; Table S2). Knowledge of the correct mortality rate of 1 in 3 remained low and unchanged (Figure S1). In 2025, 82% respondents were unaware of PSS, and only 6% reported knowing someone who had experienced it (Figure 1 and Table S2).

Sepsis awareness increased among both men and women. However, women consistently reported significantly higher overall sepsis awareness (46%-75%) than men (34%-65%), all p<.001. Awareness was higher among women showed higher recognition of all high-risk groups for sepsis than men (36% vs 26%), though knowledge of the correct sepsis mortality rate, PSS and sepsis-COVID-19 link did not differ by gender. Respondents aged ≥50 years reported the highest recognition of at-risk groups for sepsis (38%) but the lowest PSS awareness (13%; Table S3).

All Australian States and Territories showed sustained increased, except in Western Australia, where awareness peaked in 2022, but declined in 2025. Queensland recorded the highest awareness in 2016 (43%) and 2020 (67%), which was surpassed by Western Australia in 2022 (72%), while in 2025 group of smaller jurisdictions (Tasmania/ACT/Norther Territory) recorded the highest awareness of 84%. Overall sepsis awareness differed significantly (p<.05) across states in all surveys, except 2016. Knowledge of high risk for sepsis of pregnancy and postpartum was uniformly low (4%-10%). Sepsis awareness increased from 38% to 64% among capital city residents and 43% to 80% among non-capital residents, with the difference being statistically significant in 2022 and 2025 (p<.001). Except for awareness of the sepsis-COVID-19 link and sepsis media coverage, non-capital city residents reported higher sepsis awareness (Table S4).

Married respondents and parents of children under 18 years reported higher awareness of PSS and media coverage social media being a common source of sepsis information (Table S5). Awareness increased across all household income groups with no consistent trend in low versus high income groups (Table S6). Among various generations, millennials demonstrated the highest knowledge of PSS (26%), sepsis-COVID-19 link (27% in 2025) and media coverage (42% in 2022), while Baby Boomers recorded the highest knowledge of high-risk groups for sepsis (40%) (Table S7).

In 2025 survey, common information sources were social circle (family/friends/colleagues, 34%), traditional media such as TV and radio (27%), and social media (16%) (Table S8). Among those with lived experience of sepsis, 42% were family/friend of a sepsis survivor (Figure S4). Sepsis Australia and ACSQHC accounts were the most frequently cited social media sources (13% each) (Figure S5). More women received sepsis information through lived experience or their social circle, whereas men cited traditional media as their common source of sepsis information. Social media use as an information source was more common in younger respondents and younger generations. Sources of sepsis information were generally similar in other subgroups (Table S8). Only 10% respondents rated federal or state government support for sepsis patients and their families as adequate, while 46% indicated that additional government support is needed, a finding consistent across subgroups (Table S9).

## Discussion

### Summary of key findings

This longitudinal analysis of four nationally representative surveys demonstrates a significant increase in self-reported public awareness of sepsis in Australia since 2016, yet it remains lower than awareness of other common conditions. Activities undertaken by Sepsis Australia and other organisations including the ACSQHC during this period may have contributed to this improvement, although a causal relationship is not proven. Importantly, improvement in overall awareness did not translate into improved knowledge about sepsis, reflected by continued low knowledge of sepsis signs and symptoms and mortality rate as well as limited knowledge of long-term effects of sepsis and population groups most at risk.

Women, older respondents, non-capital residents, parents of children <18 years, and unemployed respondents reported higher sepsis awareness. Awareness of the sepsis-COVID-19 link peaked in 2022, coinciding with the pandemic. Most respondents perceived government support for sepsis as inadequate.

### Comparison with previous studies

In 2025, 70% of Australian participants were aware of sepsis, similar to Canadian (61%) and the Sepsis Alliance United States (65%) surveys (12) and a previous survey among Australian parents (61.6%). (14) While lower than the 88.6% reported in a German survey of elderly individuals, 2025 sepsis awareness in our study is higher than several other countries, including Ireland, France, Singapore, Spain, Brazil, and Sweden. (15, 16) Knowledge of fever as a sepsis symptom was consistent with findings from a recent scoping review. (16) Gender differences in awareness mimics that of other conditions, such as, higher awareness mental health literacy among women. (17)

### Strengths and limitations

Strengths of this study include comprehensive evaluation of trends in public sepsis awareness in Australia across four nationally representative surveys. Moreover, survey data were weighted by age, gender, and region to reflect the latest ABS population estimates. Inclusion of awareness of the longer-term effects of sepsis in survivors and perception of government support adds important context to public understanding. Limitations include potential biases inherent to survey research, (18) inability to infer causality between awareness activities and improvement in sepsis awareness, and changes in survey questions across four surveys that limited trend analysis for some parameters.

### Implications and next steps

Findings from this study highlight priorities for improving public sepsis awareness in Australia. Between 2019 and 2025, two tranches of federal government funding totalling AU$3.6 million supported the National Sepsis Program with sepsis awareness initiatives and activities undertaken by Sepsis Australia forming a minor component of that program. (19) Despite these efforts sepsis awareness remains lower than other major health conditions, and evidence suggest that health awareness declines without sustained advocacy. Stable, multi-year funding support is therefore likely needed to improve sepsis awareness further. (20) When asked, survey respondents overwhelming supported greater government action.

Globally, paid media channels have played a key role in successful health campaigns, including those for breast cancer and coronary heart disease. (21–23) Sepsis awareness campaigns and initiatives using social media, earned and commercial media, public transport and ambulance advertising, have yielded positive results in the United Kingdom. (24) Australia should implement a similar comprehensive annual national sepsis awareness campaign, as recommended by the sepsis national awareness campaign scoping review undertaken by Sepsis Australia and the ACSQHC. (25) Targeted strategies are needed to for groups with disproportionately low sepsis awareness, including men and First Nation people. Integration into existing initiatives such as, ‘Closing the Gap’ (26) for First Nations people, and antenatal care for pregnant and postpartum women as a high-risk group for sepsis, may enhance reach. Lower sepsis awareness among men warrants tailored approaches, given they experience higher sepsis mortality and poorer outcomes. (27)

Sepsis awareness peaked in different states and territories in different years possibly reflecting inconsistent awareness campaigns and localised media coverage. (28, 29) Evidence from a recent German survey suggests that sepsis-related emergencies are perceived as less urgent than other emergencies such as stroke or myocardial infarction, potentially delaying care-seeking. (30) Future Australian sepsis awareness campaigns should therefore emphasise on improving public understanding of urgency of sepsis and the importance of seeking timely medical care.

## Conclusion

Public awareness of sepsis in Australia has improved in recent years, likely reflecting the impact of national initiatives by Sepsis Australia, the ACSQHC and other organisations. Crucial gaps persist among men, younger adults, capital cities residents, First Nations people, parents and unemployed individuals, and in knowledge of signs and symptoms, mortality rate, at risk group and PSS. A comprehensive, targeted national awareness campaign, supported by sustained funding, is warranted to close these gaps and reduce preventable deaths and disabilities.

## Data Availability

All data produced in the present study are available upon reasonable request to the authors

