## Supplementary File 1 for "Trends in public awareness of sepsis in Australia from 2016 to 2025: a longitudinal analysis of four national surveys (Preliminary results)"

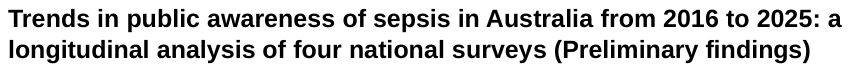

**Supplementary File 1: Survey questions**

| Question number | **Survey question** | **2016** | **2020** | **2022** | **2025** |
| --- | --- | --- | --- | --- | --- |
| A1 | Location | Yes | Yes | Yes | Yes |
| A2 | Gender | Yes | Yes | Yes | Yes |
| A3 | Age | Yes | Yes | Yes | Yes |
| A4 | First Nation status | No | No | Yes | Yes |
| B1 | Thinking now about health. Which of these medical conditions have you heard of before today? (includes Sepsis) Please select all that apply. | Yes | Yes | Yes | Yes |
| B2 | Thinking now just about the condition known as sepsis. To the best of your knowledge, what are the main symptoms of sepsis? Please be specific. (open-ended) | Yes | Yes | Yes | Yes |
| B3 | And to the best of your knowledge, what is the main cause of sepsis? Please be specific. (open-ended) | Yes | Yes | Yes | Yes |
| B4 | Do you know someone who has had sepsis when suffering from an infection? | Yes | Yes | Yes | Yes |
| B5 | To the best of your knowledge, what proportion of people diagnosed with sepsis will die from it? Please select one option only. | Yes | Yes | Yes | Yes |
| B6 | Before taking this survey, to what extent were you aware of the link between COVID-19 and sepsis? | No | Yes | Yes | Yes |
| C1 | In the last 12 months, have you heard/seen anything in the news or other sources about Australians who have had or died from sepsis? | No | No | Yes | Yes |
| C1_ | How did you hear about sepsis? | No | No | No | Yes |
| C1a | You mentioned you have lived experience of sepsis. Which of the following best describes your experience? | No | No | No | Yes |
| C1b | Where on social media did you hear about sepsis? | No | No | No | Yes |
| D1 | Many sepsis survivors experience a significant burden of physical and psychological health problems, a reduced health-related quality of life and increased risk of death in subsequent months and years. This condition is known as post-sepsis Syndrome before today, were you aware of post-sepsis syndrome? | No | No | No | Yes |
| D2 | Which of the following groups of people do you think are at greater risk of getting sepsis? Please select all that apply | No | No | No | Yes |
| D3 | Please indicate your views about the level of Federal and State Government support (e.g. funding raising sepsis awareness and education, clinical care or research and awareness activities) for Sepsis. | | | | |
| D3_1 | Federal Government | No | No | No | Yes |
| D3_2 | State Government | No | No | No | Yes |
