## Supplementary File 2 for "Trends in public awareness of sepsis in Australia from 2016 to 2025: a longitudinal analysis of four national surveys (Preliminary results)"

**Table S1 Reported symptoms and causes of sepsis**

|  |  | 2016  (N=1000) | 2020  (N=1006) | 2022  (N=1131) | 2025  (N=1232) |
| --- | --- | --- | --- | --- | --- |
| Symptoms of sepsis | Blood poisoning/ infection/ disease | 8% | 11% | 8% | 11% |
|  | Fever/ high temperature/ chill/ sweating | 14% | 23% | 28% | 36% |
|  | Bacterial Infection | NR | 1% | 0% | 1% |
|  | Infection of a wound | NR | 1% | 1% | 2% |
|  | Other infection | 6% | 9% | 9% | 8% |
|  | Headache | NR | 1% | 2% | 2% |
|  | Nausea/ vomiting | NR | 6% | 6% | 7% |
|  | Pain | NR | 7% | 5% | 12% |
|  | Swelling / inflammation/ redness | NR | 8% | 7% | 10% |
|  | Decreased urination | NR | NR | 1% | 2% |
|  | Rapid Pulse/ heart rate | NR | 4% | 6% | 9% |
|  | Rapid breathing/ breathing changes | NR | 6% | 7% | 10% |
|  | Unconsciousness/ coma | NR | 1% | 2% | 1% |
|  | Death | NR | 2% | 1% | 2% |
|  | Lethargy | NR | NR | 5% | 7% |
|  | Diarrhoea | NR | 1% | 0% | 1% |
|  | Confusion/ delirium | NR | 7% | 8% | 8% |
|  | Changed blood pressure | NR | 4% | 5% | 3% |
|  | Organ failure | NR | 3% | 2% | 5% |
|  | Feeling unwell | NR | 3% | 3% | 6% |
|  | Pus/ sores | NR | 1% | 1% | 1% |
|  | Skin colour changes/ issues | NR | NR | 3% | NR |
|  | Other | NR | 9% | 12% | 17% |
|  | Don't know | 9% | 13% | 11% | 8% |
| Causes of sepsis | Blood poisoning/ bacterial infection/ disease | 7% | 8% | 10% | 11% |
|  | Bacterial Infection | 3% | 4% | 9% | 6% |
|  | (Bacterial) Infection of a wound | NR | NR | 2% | 3% |
|  | Other infection | 11% | 20% | 24% | 32% |
|  | Untreated/ complication of an infection | NR | 2% | 2% | 3% |
|  | Contamination of a wound | NR | 7% | 3% | 1% |
|  | Germs/ toxins | NR | 1% | 2% | 1% |
|  | Immune Response | NR | 3% | 2% | 6% |
|  | Pneumonia | NR | NR | 2% | NR |
|  | Low/high blood pressure | NR | NR | 0% | NR |
|  | Virus | NR | NR | 1% | NR |
|  | COVID-19 | NR | NR | 1% | NR |
|  | Other | NR | 6% | 9% | 13% |
|  | Don't know | 10% | 15% | 11% | 10% |
| Not heard of sepsis | | 60% | 42% | 40% | 31% |

NR: Not reported. Note: Surveyed samples were weighted as per latest population estimates from the Australian Bureau of Statistics as follows: 2016 = 18,228; 2020 = 20,076; 2022 = 20,076; 2025 = 21,651

**Table S2 Other aspects of sepsis awareness in total survey population**

|  | Year | N | Awareness |
| --- | --- | --- | --- |
| Knew correct sepsis mortality rate (1 in 3) | 2016 | 1000 | 4% |
|  | 2020 | 1006 | 4% |
|  | 2022 | 1131 | 6% |
|  | 2025 | 1232 | 5% |
| Knew someone with sepsis | 2016 | 1000 | 12% |
|  | 2020 | 1006 | 16% |
|  | 2022 | 1131 | 22% |
|  | 2025 | 1232 | 22% |
| Knew link between COVID-19 and sepsis | 2020 | 1006 | 17% |
|  | 2022 | 1131 | 21% |
|  | 2025 | 1232 | 17% |
| Knew sepsis media coverage in previous 12 months | 2022 | 1131 | 30% |
|  | 2025 | 1232 | 29% |
| Post-Sepsis Syndrome awareness |  | 1232 |  |
| Yes, and know someone who has experienced it  Yes, but don't know anyone who has experienced it  Not aware | 2025  2025  2025 |  | 6%  12%  82% |
| High-risk groups |  | 1232 |  |
| Newborn & Youth | 2025 |  | 20% |
| Older people | 2025 |  | 41% |
| Aboriginal & Torres Strait Islander Peoples | 2025 |  | 14% |
| People with Complex Health Problems | 2025 |  | 35% |
| People with COVID-19 | 2025 |  | 34% |
| People with poor immune systems | 2025 |  | 40% |
| Currently pregnant or post pregnancy | 2025 |  | 8% |
| Has cancer and undergoing chemotherapy | 2025 |  | 20% |
| People with burns, wounds and injuries | 2025 |  | 22% |
| People previously diagnosed with Sepsis | 2025 |  | 25% |
| All of the above | 2025 |  | 31% |

Note: Surveyed samples were weighted as per latest population estimates from the Australian Bureau of Statistics as follows: 2016 = 18,228; 2020 = 20,076; 2022 = 20,076; 2025 = 21,651

**Table S3 Sepsis awareness by gender and age groups**

|  | Year | N | Gender | | Age groups (years) | | | |
| --- | --- | --- | --- | --- | --- | --- | --- | --- |
|  |  |  | Men | Women | 18-24 | 25-34 | 35-49 | 50+ |
| Awareness of PSS | 2025 | 1232 | 18% | 18% | 19% | 27% | 22% | 13% |
| Knew correct Sepsis Mortality Rate* | 2016 | 1000 | 4% | 3% | 5% | 6% | 3% | 3% |
|  | 2020 | 1006 | 3% | 5% | 3% | 4% | 4% | 4% |
|  | 2022 | 1131 | 6% | 6% | 9% | 6% | 6% | 5% |
|  | 2025 | 1232 | 6% | 5% | 5% | 9% | 4% | 5% |
| Knew someone with sepsis | 2016 | 1000 | 10% | 14% | 10% | 16% | 9% | 13% |
|  | 2020 | 1006 | 13% | 19% | 9% | 16% | 20% | 16% |
|  | 2022 | 1131 | 20% | 24% | 15% | 26% | 25% | 20% |
|  | 2025 | 1232 | 17% | 26% | 12% | 20% | 2% | 26% |
| Link between sepsis and COVID-19 | 2020 | 1006 | 18% | 16% | 15% | 26% | 18% | 14% |
|  | 2022 | 1131 | 24% | 19% | 22% | 34% | 31% | 10% |
|  | 2025 | 1232 | 18% | 16% | 15% | 26% | 23% | 11% |
| Exposed to sepsis media coverage in past 12 months | 2022 | 1131 | 32% | 29% | 25% | 42% | 41% | 21% |
|  | 2025 | 1232 | 29% | 29% | 33% | 40% | 32% | 23% |
| Correct knowledge of all at-risk groups for sepsis | 2025 | 1232 | 26% | 36% | 34% | 20% | 26% | 38% |

PSS: Post-sepsis syndrome. *Mortality rate: proportion of people diagnosed with sepsis who die, respondents who selected sepsis mortality rate as 1 in 3.

Note: Surveyed samples were weighted as per latest population estimates from the Australian Bureau of Statistics as follows: 2016 = 18,228; 2020 = 20,076; 2022 = 20,076; 2025 = 21,651

**Table S4 Sepsis awareness as per geographical location**

|  | Year | N | NSW | VIC | QLD | SA | WA | TAS/NT/ACT | Capital Cities | Non-capital cities |
| --- | --- | --- | --- | --- | --- | --- | --- | --- | --- | --- |
| Knew correct sepsis mortality rate | 2016 | 1000 | 3% | 5% | 4% | 4% | 2% | NR | 3% | 4% |
|  | 2020 | 1006 | 3% | 5% | 3% | 7% | 2% | 7% | 4% | 4% |
|  | 2022 | 1131 | 6% | 5% | 4% | 4% | 7% | 9% | 5% | 7% |
|  | 2025 | 1232 | 6% | 6% | 5% | 4% | 5% | 5% | 5% | 6% |
| Knew someone with sepsis | 2016 | 1000 | 11% | 12% | 12% | 9% | 16% | NR | 11% | 12% |
|  | 2020 | 1006 | 16% | 17% | 16% | 20% | 12% | 16% | 13% | 22% |
|  | 2022 | 1131 | 24% | 18% | 22% | 27% | 17% | 29% | 20% | 24% |
|  | 2025 | 1232 | 19% | 21% | 22% | 27% | 26% | 24% | 21% | 23% |
| Knew about link between COVID-19 and sepsis | 2020 | 1006 | 16% | 16% | 21% | 20% | 16% | 11% | 18% | 16% |
|  | 2022 | 1131 | 22% | 20% | 22% | 28% | 15% | 21% | 24% | 16% |
|  | 2025 | 1232 | 18% | 18% | 13% | 17% | 21% | 10% | 21% | 10% |
| Knew sepsis media coverage in previous 12 months | 2022 | 1131 | 30% | 29% | 34% | 31% | 30% | 32% | 33% | 26% |
|  | 2025 | 1232 | 11% | 8% | 9% | 7% | 16% | 7% | 11% | 8% |
| Post sepsis Syndrome awareness | 2025 | 1232 | 18% | 21% | 14% | 22% | 25% | 13% | 22% | 13% |
| High-risk groups |  | 1232 |  |  |  |  |  |  |  |  |
| Newborn & Youth | 2025 |  | 22% | 22% | 16% | 19% | 16% | 25% | 21% | 19% |
| Older people | 2025 |  | 41% | 43% | 42% | 37% | 33% | 54% | 41% | 42% |
| Aboriginal & Torres Strait Islander Peoples | 2025 |  | 17% | 12% | 16% | 12% | 5% | 19% | 13% | 16% |
| People with Complex Health Problems | 2025 |  | 36% | 38% | 33% | 28% | 28% | 49% | 36% | 33% |
| People with COVID-19 | 2025 |  | 34% | 35% | 32% | 28% | 35% | 42% | 34% | 33% |
| People with poor immune systems | 2025 |  | 40% | 40% | 41% | 40% | 32% | 48% | 41% | 37% |
| Currently Pregnant or post pregnancy | 2025 |  | 10% | 8% | 4% | 7% | 6% | 17% | 10% | 6% |
| Has cancer and undergoing chemotherapy | 2025 |  | 19% | 21% | 19% | 21% | 15% | 26% | 20% | 19% |
| People with burns, wounds and injuries | 2025 |  | 22% | 22% | 22% | 23% | 17% | 37% | 22% | 22% |
| People previously diagnosed with Sepsis | 2025 |  | 25% | 27% | 23% | 25% | 22% | 36% | 26% | 25% |
| All of the above | 2025 |  | 30% | 29% | 33% | 32% | 37% | 29% | 29% | 34% |

ACT: Australian Capital Territory, NSW: New South Wales, NR: Not reported; NT: Northern Territory, VIC: Victora, SA: South Australia, QLD: Queensland, TAS: Tasmania, WA: Western Australia. Mortality rate: defined as the proportion of people diagnosed with sepsis who die.

Note: Surveyed samples were weighted as per latest population estimates from the Australian Bureau of Statistics as follows: 2016 = 18,228; 2020 = 20,076; 2022 = 20,076; 2025 = 21,651

**Table S5 Sepsis awareness as per marital status and having child at home**

|  | Year | N | Marital status | | Children (<18 years) at home | |
| --- | --- | --- | --- | --- | --- | --- |
|  |  |  | Married | Not married | Yes | No |
| Knew correct sepsis mortality rate (1 in 3) | 2016 | 1000 | 3% | 4% | 5% | 3% |
|  | 2020 | 1006 | 4% | 3% | 5% | 3% |
|  | 2022 | 1131 | 6% | 6% | 4% | 6% |
|  | 2025 | 1232 | 6% | 5% | 6% | 5% |
| Knew someone with sepsis | 2016 | 1000 | 14% | 8% | 12% | 12% |
|  | 2020 | 1006 | 17% | 15% | 17% | 16% |
|  | 2022 | 1131 | 23% | 19% | 21% | 22% |
|  | 2025 | 1232 | 24% | 19% | 23% | 21% |
| Knew link between COVID-19 and sepsis | 2020 | 1006 | 17% | 18% | 24% | 14% |
|  | 2022 | 1131 | 21% | 22% | 31% | 15% |
|  | 2025 | 1232 | 19% | 14% | 27% | 13% |
| Knew sepsis media coverage in previous 12 months | 2022 | 1131 | 34% | 26% | 41% | 24% |
|  | 2025 | 1232 | 11% | 9% | 13% | 9% |
| Post Sepsis Syndrome Awareness | 2025 | 1232 | 23% | 13% | 28% | 14% |
| High-risk groups |  | 1232 |  |  |  |  |
| Newborn & Youth | 2025 |  | 20% | 20% | 23% | 19% |
| Older people | 2025 |  | 40% | 42% | 41% | 42% |
| Aboriginal & Torres Strait Islander Peoples | 2025 |  | 14% | 13% | 12% | 16% |
| People with Complex Health Problems | 2025 |  | 34% | 37% | 36% | 36% |
| People with COVID-19 | 2025 |  | 35% | 33% | 38% | 33% |
| People with poor immune systems | 2025 |  | 39% | 41% | 42% | 39% |
| Currently Pregnant or post pregnancy | 2025 |  | 8% | 8% | 11% | 7% |
| Has cancer and undergoing chemotherapy | 2025 |  | 19% | 20% | 17% | 21% |
| People with burns, wounds and injuries | 2025 |  | 20% | 25% | 19% | 25% |
| People previously diagnosed with Sepsis | 2025 |  | 24% | 27% | 26% | 25% |
| All of the above | 2025 |  | 31% | 32% | 28% | 32% |

Mortality rate: defined as the proportion of people diagnosed with sepsis who die.

Note: Surveyed samples were weighted as per latest population estimates from the Australian Bureau of Statistics as follows: 2016 = 18,228; 2020 = 20,076; 2022 = 20,076; 2025 = 21,651

**Table S6 Sepsis awareness as per household income and employment status**

|  | Year | N | <$50K | $50K- $99K | ≥$100K | Full-time | Part-time | Not working |
| --- | --- | --- | --- | --- | --- | --- | --- | --- |
| Knew correct sepsis mortality rate (1 in 3) | 2016 | 1000 | 4% | 4% | 3% | 5% | 3% | 2% |
|  | 2020 | 1006 | 3% | 4% | 4% | 5% | 4% | 4% |
|  | 2022 | 1131 | 4% | 5% | 7% | 6% | 8% | 5% |
|  | 2025 | 1232 | 5% | 4% | 7% | 7% | 2% | 7% |
| Knew someone with sepsis | 2016 | 1000 | 11% | 12% | 12% | 12% | 9% | 13% |
|  | 2020 | 1006 | 18% | 15% | 18% | 17% | 19% | 14% |
|  | 2022 | 1131 | 18% | 26% | 22% | 24% | 25% | 17% |
|  | 2025 | 1232 | 21% | 20% | - | 20% | 23% | 19% |
| Knew about link between COVID-19 and sepsis | 2020 | 1006 | 16% | 18% | 19% | 24% | 18% | 12% |
|  | 2022 | 1131 | 19% | 20% | 27% | 32% | 19% | 12% |
|  | 2025 | 1232 | 11% | 20% | - | 26% | 11% | 8% |
| Knew sepsis media coverage in previous 12 months | 2022 | 1131 | 22% | 35% | 38% | 43% | 28% | 18% |
|  | 2025 | 1232 | 8% | 11% | - | 13% | 8% | 6% |
| Post Sepsis Syndrome Awareness | 2025 | 1232 | 16% | 18% | 46% | 25% | 15% | 21% |
| High-risk groups |  | 1232 |  |  |  |  |  |  |
| Newborn & Youth | 2025 |  | 20% | 18% | 42% | 22% | 20% | 38% |
| Older people | 2025 |  | 41% | 43% | 85% | 44% | 39% | 77% |
| Aboriginal & Torres Strait Islander Peoples | 2025 |  | 15% | 17% | 27% | 13% | 14% | 27% |
| People with Complex Health Problems | 2025 |  | 34% | 38% | 75% | 40% | 28% | 66% |
| People with COVID-19 | 2025 |  | 36% | 36% | 65% | 38% | 35% | 55% |
| People with poor immune systems | 2025 |  | 40% | 40% | 87% | 43% | 37% | 75% |
| Currently Pregnant or post pregnancy | 2025 |  | 8% | 7% | 21% | 9% | 8% | 15% |
| Has cancer and undergoing chemotherapy | 2025 |  | 20% | 24% | 36% | 20% | 18% | 43% |
| People with burns, wounds and injuries | 2025 |  | 25% | 22% | 45% | 21% | 18% | 52% |
| People previously diagnosed with Sepsis | 2025 |  | 24% | 26% | 53% | 27% | 24% | 49% |
| All of the above | 2025 |  | 35% | 28% | 60% | 24% | 36% | 74% |

Mortality rate: defined as the proportion of people diagnosed with sepsis who die. Note: $ indicates Australia dollars.

Note: Surveyed samples were weighted as per latest population estimates from the Australian Bureau of Statistics as follows: 2016 = 18,228; 2020 = 20,076; 2022 = 20,076; 2025 = 21,651

**Table S7 Sepsis awareness across various generations**

|  | Year | N | Next Gen (18 to 34 yrs) | GenZ (1997- 2009) | Millennials (1981- 1996) | GenX (1965-1980) | Baby Boomer (1946-1964) | Silent (1918- 1945) |
| --- | --- | --- | --- | --- | --- | --- | --- | --- |
| Knew someone with sepsis | 2020 | 1000 | 22% | 14% | 23% | 18% | 13% | 11% |
|  | 2022 | 1006 | 30% | 24% | 34% | 23% | 7% | 12% |
|  | 2025 | 1131 | 17% | 15% | 19% | 25% | 26% | 23% |
| Knew correct sepsis mortality rate (1 in 3) | 2020 | 1232 | 4% | 1% | 4% | 5% | 4% | 2% |
|  | 2022 | 1000 | 7% | 9% | 5% | 5% | 6% | 1% |
|  | 2025 | 1006 | 7% | 6% | 6% | 5% | 5% | - |
| Knew about link between COVID-19 and sepsis | 2020 | 1131 | 13% | 6% | 16% | 21% | 15% | 14% |
|  | 2022 | 1232 | 22% | 16% | 24% | 25% | 21% | 9% |
|  | 2025 | 1006 | 22% | 19% | 27% | 11% | 9% | 15% |
| Knew sepsis media coverage in previous 12 months | 2022 | 1131 | 35% | 24% | 42% | 37% | 19% | 12% |
|  | 2025 | 1232 | 11% | 10% | 12% | 7% | 10% | 13% |
| Post Sepsis Syndrome awareness | 2025 | 1131 | 24% | 21% | 26% | 12% | 13% | 18% |
| **High-risk groups** |  | 1232 |  |  |  |  |  |  |
| Newborn & Youth | 2025 |  | 23% | 27% | 20% | 20% | 18% | 11% |
| Older people | 2025 |  | 42% | 39% | 42% | 43% | 40% | 33% |
| Aboriginal & Torres Strait Islander Peoples | 2025 |  | 9% | 11% | 11% | 19% | 16% | 15% |
| People with Complex Health Problems | 2025 |  | 38% | 38% | 38% | 32% | 34% | 26% |
| People with COVID-19 | 2025 |  | 40% | 37% | 37% | 32% | 31% | 26% |
| People with poor immune systems | 2025 |  | 43% | 41% | 43% | 38% | 37% | 35% |
| Currently Pregnant or post pregnancy | 2025 |  | 12% | 16% | 11% | 5% | 4% | 2% |
| Has cancer and undergoing chemotherapy | 2025 |  | 19% | 19% | 19% | 23% | 19% | 16% |
| People with burns, wounds and injuries | 2025 |  | 21% | 19% | 19% | 26% | 25% | 25% |
| People previously diagnosed with Sepsis | 2025 |  | 29% | 27% | 27% | 24% | 25% | 18% |
| All of the above | 2025 |  | 25% | 30% | 24% | 32% | 40% | 28% |

Mortality rate: defined as the proportion of people diagnosed with sepsis who die.

Note: 18 to 34 (Next Gen) was excluded from the comparison of various generation due to overlap with GenZ and millennials. Surveyed samples were weighted as per latest population estimates from the Australian Bureau of Statistics as follows: 2016 = 18,228; 2020 = 20,076; 2022 = 20,076; 2025 = 21,651

**Table S8: Sources of sepsis information in various subgroups (2025 survey)**

| Category | Lived experience of sepsis* | Heard from family, friends, colleagues | GP | Social media | Traditional media** | Sepsis Australia website | ACSQHC website | State/Territory Sepsis Program site | Sepsis programs in other countries websites | Other | Can’t recall/Don’t know |
| --- | --- | --- | --- | --- | --- | --- | --- | --- | --- | --- | --- |
| **Total survey population (N = 866)** | 13% | 34% | 5% | 16% | 27% | 2% | 3% | 2% | 1% | 12% | 22% |
| **Gender** |  |  |  |  |  |  |  |  |  |  |  |
| Male | 9% | 27% | 7% | 18% | 30% | 2% | 5% | 3% | 2% | 9% | 27% |
| Females | 16% | 39% | 4% | 13% | 24% | 2% | 2% | 1% | 1% | 13% | 17% |
| **Age groups** |  |  |  |  |  |  |  |  |  |  |  |
| 18 to 24 | 1% | 34% | 4% | 34% | 33% | 2% | 6% | 10% | 3% | 10% | 16% |
| 25 to 34 | 12% | 37% | 2% | 32% | 24% | 4% | 8% | 4% | 3% | 10% | 17% |
| 35 to 49 | 11% | 30% | 7% | 19% | 26% | 3% | 3% | 2% | 3% | 13% | 19% |
| 50 or more | 16% | 34% | 6% | 7% | 27% | 1% | 2% | 0% | 0% | 12% | 25% |
| **Place of residence** | | | | | | | | | | | |
| NSW | 13% | 34% | 6% | 17% | 19% | 2% | 2% | 1% | 1% | 9% | 19% |
| VIC | 12% | 32% | 5% | 18% | 26% | 1% | 3% | 1% | 0% | 13% | 24% |
| QLD | 13% | 33% | 7% | 13% | 26% | 2% | 3% | 0% | 0% | 11% | 21% |
| SA | 16% | 33% | 5% | 11% | 26% | 2% | 3% | 8% | 0% | 17% | 19% |
| WA | 8% | 35% | 6% | 17% | 24% | 5% | 4% | 6% | 2% | 9% | 26% |
| TAS/NT/ACT | 16% | 35% | 5% | 9% | 22% | 0% | 2% | 1% | 0% | 18% | 25% |
| Capital city | 12% | 36% | 6% | 22% | 28% | 3% | 4% | 3% | 2% | 12% | 17% |
| Non-capital city | 14% | 30% | 4% | 7% | 25% | 0% | 2% | 0% | 1% | 11% | 28% |
| **Marital Status** |  |  |  |  |  |  |  |  |  |  |  |
| Married | 14% | 34% | 6% | 17% | 24% | 3% | 4% | 2% | 2% | 10% | 19% |
| Not Married | 11% | 35% | 5% | 14% | 24% | 0% | 2% | 1% | 0% | 13% | 25% |
| **Children <18 yr at home** | | | | | | | | | | | |
| - Yes | 16% | 31% | 4% | 29% | 27% | 4% | 7% | 5% | 4% | 9% | 16% |
| - No | 12% | 34% | 6% | 20% | 27% | 1% | 6% | 1% | 0% | 12% | 24% |
| **Household income** | | | | | | | | | | | |
| < $50K | 14% | 30% | 5% | 11% | 25% | 1% | 2% | 0% | 1% | 9% | 28% |
| $50K - $99K | 14% | 36% | 5% | 15% | 27% | 2% | 3% | 3% | 1% | 9% | 20% |
| ≥$100K | 14% | 32% | 5% | 22% | 28% | 3% | 6% | 3% | 2% | 11% | 16% |
| **Working status** | | | | | | | | | | | |
| Full Time | 12% | 36% | 6% | 23% | 26% | 4% | 6% | 4% | 1% | 10% | 18% |
| Part Time | 13% | 36% | 3% | 15% | 26% | 2% | 2% | 1% | 2% | 12% | 20% |
| Working | 12% | 33% | 5% | 20% | 26% | 2% | 5% | 3% | 2% | 11% | 18% |
| **Generations** |  |  |  |  |  |  |  |  |  |  |  |
| 18 to 34 (Next Gen) | 9% | 36% | 3% | 33% | 28% | 3% | 7% | 6% | 3% | 10% | 17% |
| Gen Z (1997 - 2009) | 6% | 33% | 5% | 31% | 27% | 2% | 5% | 7% | 3% | 12% | 18% |
| Millennials (1981 - 1996) | 12% | 33% | 5% | 28% | 27% | 4% | 7% | 4% | 3% | 11% | 16% |
| Gen X (1965 - 1980) | 13% | 37% | 6% | 9% | 21% | 0% | 1% | 0% | 0% | 14% | 22% |
| Baby Boomer (1946 - 1964) | 16% | 31% | 6% | 7% | 30% | 1% | 2% | 0% | 0% | 11% | 25% |
| Silent (1918 - 1945) | 13% | 35% | 8% | 4% | 31% | 0% | 6% | 0% | 0% | 7% | 38% |
| **First Nation people (N = 201)** | | | | | | | | | | | |
| - Yes | 19% | 45% | 3% | 47% | 34% | 24% | 19% | 14% | 9% | 10% | 8% |
| - No | 13% | 33% | 6% | 16% | 27% | 2% | 3% | 2% | 1% | 11% | 22% |

**^*^**Survivor, caregiver, family member or bereaved; **Including TV, radio or print media. ACT: Australian Capital Territory, NT: Northern Territory; TAS: Tasmania.

Note: Surveyed sample included respondents who were aware of sepsis, and was weighted as per latest population estimates from the Australian Bureau of Statistics to 15,149.

**Table S9 Perception of government support in various subgroups (2025 survey)**

| **Category** | **Federal** | | | **State** | | |
| --- | --- | --- | --- | --- | --- | --- |
|  | Enough support | Should be doing more | Don't know | Enough support | Should be doing more | Don't know |
| Total survey population (N = 1232) | 10% | 46% | 44% | 10% | 46% | 45% |
| Men | 13% | 44% | 43% | 13% | 43% | 44% |
| Women | 7% | 49% | 44% | 6% | 48% | 46% |
| **Age groups** |  |  |  |  |  |  |
| 18-24 | 12% | 47% | 41% | 12% | 47% | 41% |
| 25-34 | 15% | 51% | 34% | 16% | 45% | 38% |
| 25-49 | 17% | 44% | 39% | 14% | 47% | 39% |
| 50 or more | 4% | 46% | 51% | 4% | 45% | 51% |
| **Location** |  |  |  |  |  |  |
| NSW | 9% | 47% | 41% | 9% | 47% | 43% |
| VIC | 11% | 46% | 41% | 10% | 47% | 43% |
| QLD | 11% | 43% | 46% | 11% | 48% | 42% |
| SA | 10% | 46% | 43% | 5% | 46% | 49% |
| WA | 8% | 46% | 46% | 5% | 45% | 46% |
| TAS/NT/ACT | 13% | 46% | 41% | 11% | 47% | 44% |
| **Capital city** |  |  |  |  |  |  |
| Yes | 13% | 46% | 40% | 13% | 45% | 42% |
| No | 5% | 47% | 49% | 4% | 47% | 49% |
| **Marital Status** |  |  |  |  |  |  |
| Married | 10% | 49% | 41% | 11% | 48% | 41% |
| Not Married | 10% | 43% | 47% | 9% | 42% | 50% |
| **Children <18 yr at home** | | | | | | |
| Yes | 16% | 53% | 32% | 15% | 52% | 33% |
| No | 8% | 43% | 49% | 8% | 42% | 50% |
| **Household income** |  |  |  |  |  |  |
| Less than A$50K | 8% | 46% | 47% | 8% | 45% | 47% |
| A$50K-$99K | 10% | 47% | 43% | 9% | 45% | 46% |
| A$100K or more | 14% | 58% | 38% | 13% | 49% | 39% |
| **Employment status** | | | | | | |
| Full Time | 16% | 49% | 35% | 15% | 45% | 37% |
| Part Time | 7% | 46% | 47% | 7% | 47% | 48% |
| Working | 13% | 48% | 40% | 12% | 46% | 41% |
| Retired | 4% | 47% | 49% | 5% | 45% | 50% |
| Other / Not Working | 6% | 40% | 54% | 4% | 39% | 55% |
| **Generations** | | | | | | |
| 18 to 34 (Next Gen) | 7% | 49% | 43% | 6% | 48% | 46% |
| Gen Z (1997 - 2009) | 12% | 47% | 41% | 12% | 47% | 42% |
| Millennials (1981 - 1996) | 15% | 51% | 34% | 15% | 48% | 37% |
| Gen X (1965 - 1980) | 17% | 44% | 39% | 16% | 44% | 40% |
| Baby Boomer (1946 - 1964) | 5% | 48% | 46% | 4% | 46% | 49% |
| Silent (1918 - 1945) | 14% | 45% | 42% | 15% | 43% | 43% |
| **First Nation people** | | | | | | |
| Yes | 25% | 60% | 15% | 27% | 59% | 15% |
| No | 10% | 46% | 44% | 9% | 45% | 46% |
| **Sepsis awareness** | | | | | | |
| Overall - Yes | 7% | 44% | 49% | 8% | 41% | 51% |
| Overall – No | 17% | 52% | 30% | 13% | 56% | 31% |
| PSS – aware | 24% | 57% | 19% | 22% | 58% | 20% |
| PSS – Not aware | 7% | 44% | 49% | 7% | 43% | 50% |
| **Sepsis media coverage in past 12 months** | | | | | | |
| Aware | 23% | `57% | 21% | 19% | 58% | 23% |
| Not aware | 5% | 42% | 52% | 6% | 40% | 53% |
| **Sepsis-COVID link** |  |  |  |  |  |  |
| Aware | 29% | 55% | 16% | 25% | 60% | 15% |
| Not aware | 6% | 45% | 49% | 7% | 43% | 51% |

A$: Australian dollar.

Surveyed sample was weighted as per latest population estimates from the Australian Bureau of Statistics to 21,651

**Figure S1 Awareness of sepsis mortality Rate**

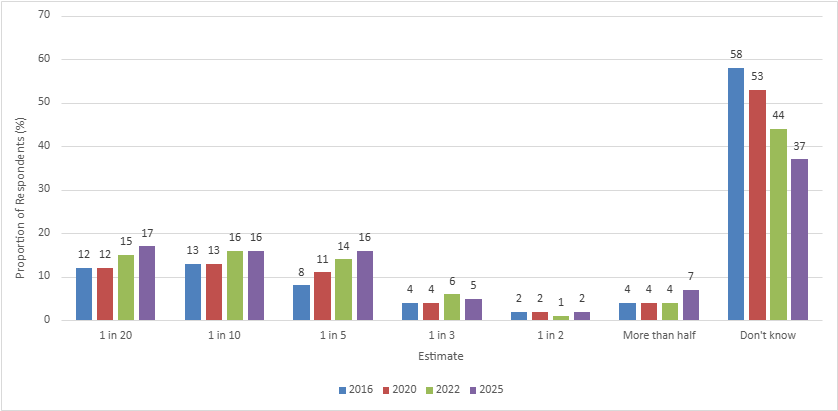

Mortality Rate: Proportion of people diagnosed with sepsis who die

**Figure S2 Sepsis awareness in First Nations people**

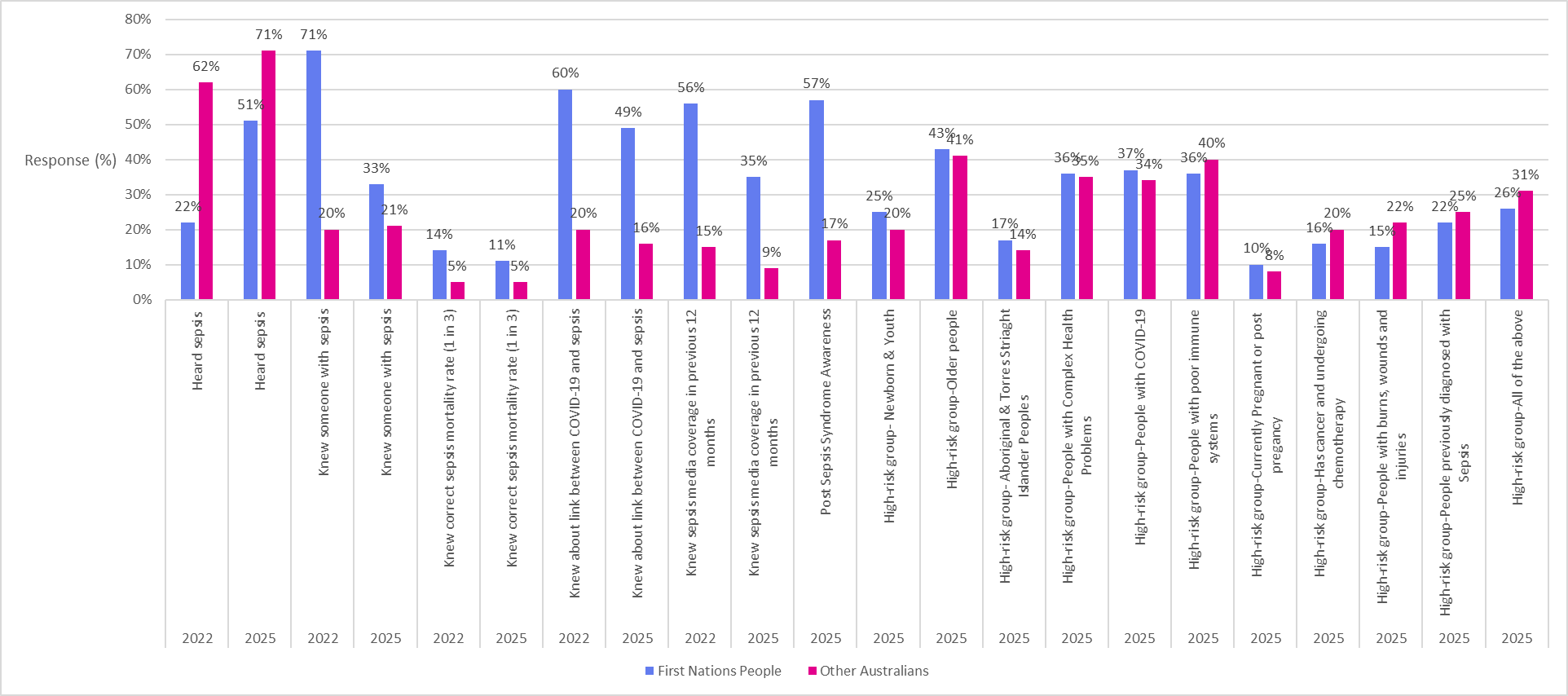

Mortality Rate: defined as the proportion of people diagnosed with sepsis who die.

Note: Sepsis awareness between First Nation and non-First nation people was statistically significant in both 2022 and 2025 (p<.001).

Number of First Nation respondents included in the 2022 and 2025 surveys were 190 and 201, respectively.

Figure S3 Awareness of link between COVID-19 and sepsis

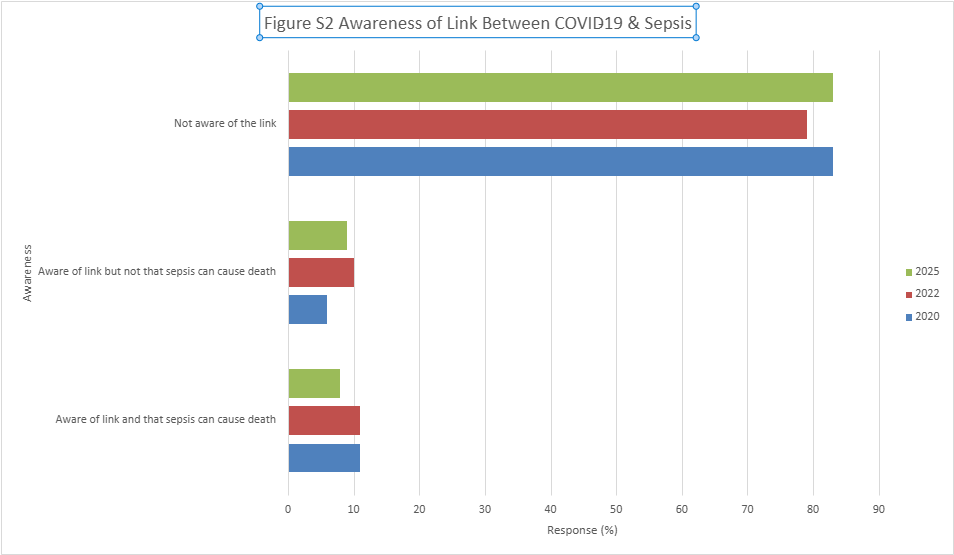

**Figure S4 Sepsis lived experience details (2025 survey)**

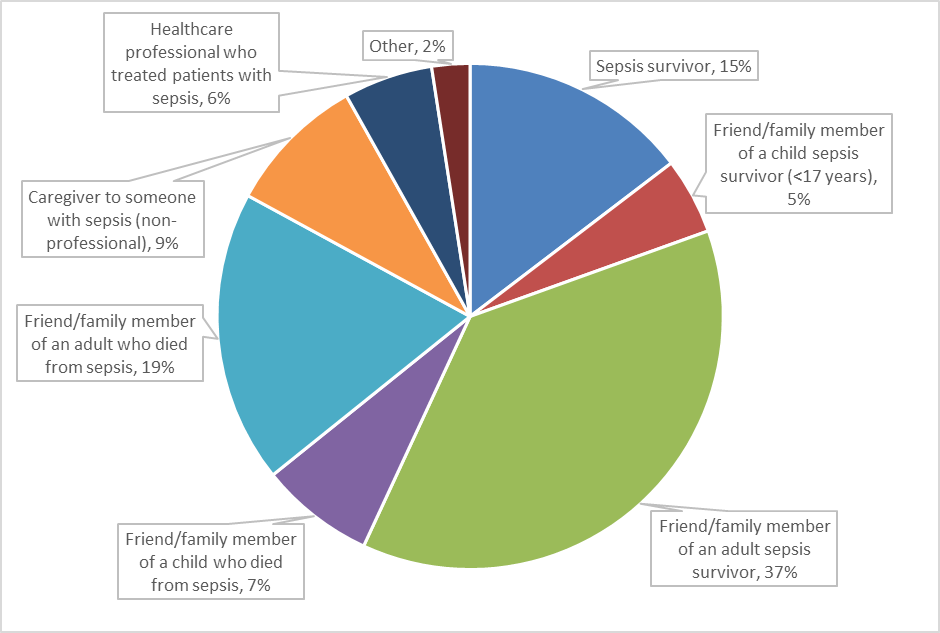

Note: Done in a subset of respondents (N=119) who responded that they have lived experienced of sepsis.

**Figure S5 Social media sources for sepsis information (2025 survey)**

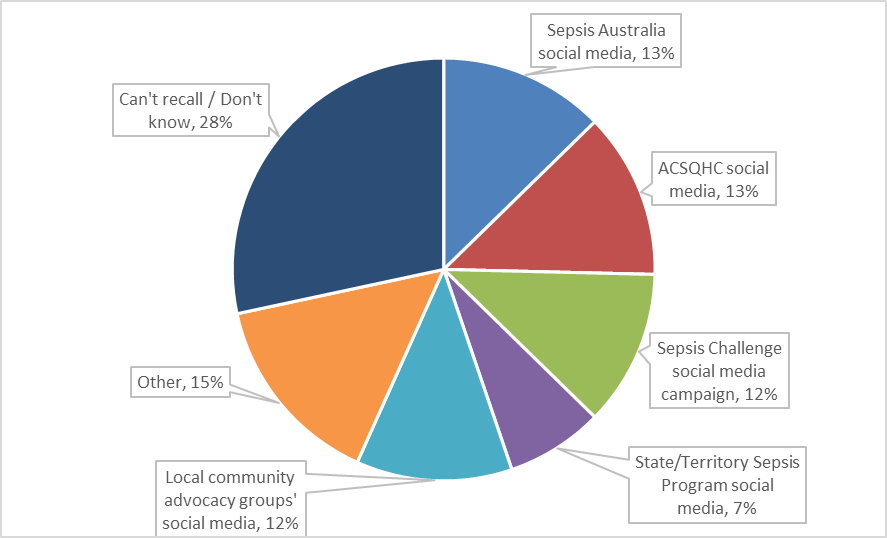

Note: Done in a subset of respondents (N=146) who responded that they use social media as the source of sepsis information.
